# An integrated framework for early detection and transmissibility assessment of emerging variants in wastewater

**DOI:** 10.1101/2025.02.18.25322479

**Authors:** Xingwen Chen, Tin Phan, Wei Lin Lee, Steven F Rhode, Samantha Brozak, Bruce Pell, Tsultrim Palden, Mats Leifels, Anna Gitter, Yang Kuang, Stefan Wuertz, Janelle Thompson, Kristina D. Mena, Eric J. Alm, Fuqing Wu

## Abstract

Tracking the emergence of new SARS-CoV-2 variants is important for a comprehensive understanding of the pandemic’s progression. However, it remains challenging due to the low variant prevalence in the early stage of an outbreak. Here, we present an integrated framework that combines three key components: early variant detection in wastewater, validation through clinical genome sequencing, and transmissibility assessment using mathematical modeling. Using the SARS-CoV-2 Omicron variant as a proof of concept, we developed a novel nested allele-specific RT-qPCR assay (NAS-PCR) for wastewater surveillance. Our framework detected Omicron in Greater Boston wastewater samples starting from September 2021, over two months before the first U.S. clinical case. We validated these findings by analyzing GISAID clinical sequence data, which revealed 172 previously unreported Omicron genomes predating its official identification in South Africa. To assess transmissibility, we developed a Susceptible-Infected-Viral load model using quantified wastewater concentrations, which estimated Omicron’s basic reproduction number (R0) between 2.36 and 3.09, showing robust consistency across varying population sizes, data points, and viral shedding rates. This integrated approach unifies molecular diagnostics, wastewater epidemiology, and mathematical modeling for comprehensive variant surveillance. Our framework provides a systematic solution for early warning and risk assessment of emerging variants, which can strengthen public health preparedness for future viral threats.

## Introduction

Since 2020, the world has witnessed waves of COVID-19 cases caused by severe acute respiratory syndrome-associated coronavirus 2 (SARS-CoV-2) and its variants including the ongoing Omicron variant. The first Omicron variant BA.1 (or B.1.1.529), was first identified in South Africa in early November 2021 ^1^. The first Omicron case in the United States was documented in San Francisco on December 1, when a fully vaccinated resident travelled back from South Africa ^2^. The Omicron variant quickly displaced Delta and became dominant, starting from less than 1% of the sequenced genomes as of December 4 to 84% by December 27 ^3^, suggesting its high transmissibility. Compared to its predecessors, the Omicron BA.1 has many more mutations (∼50 in total) including 35 in the spike gene ^4^, which leads to widespread escape from neutralizing antibodies ^4–8^. Two of the signature mutations for the Omicron variant are Q493R and Q498R, which are shared by BA.1 and BA.2, respectively. BA.2 overtook BA.1 and caused the second Omicron wave in the U.S. in March and April 2022, followed by the emergence of BA.4 and BA.5, BQ.1, XBB, and the recent EB.5 variants.

The continuous evolution of SARS-CoV-2 has underscored the importance of vigilant monitoring to track the virus and the emergence of new variants. Delayed identification has repeatedly led to catastrophic global outbreaks, as evident from previously documented outbreaks. The Zika virus epidemic 2015∼2016 in Brazil, causing congenital brain abnormalities and Guillain–Barré Syndrome, began in April 2015, with its presence dating back to as early as August 2013 ^9,10^. Similarly, the peak of the SARS-CoV (severe acute respiratory syndrome-associated coronavirus) epidemic in southeast China occurred in February 2003, though isolated cases emerged across multiple satellite cities starting from November 2002 ^11,12^. In the case of MERS-CoV (Middle East respiratory syndrome coronavirus), infections were reported for over a year with potential multiple spillovers from camels in Jordan before a major outbreak unfolded in Saudi Arabia in 2013 ^13,14^. These occurrences of transregional spread often go unnoticed, highlighting the urgent need for timely identification to implement public health measures and mitigate infections.

Beneath the urgency for timely and accurate detection lies the challenges associated with detecting viruses and their variants at low prevalence during the early stages of emergence within a population. Clinical surveillance, while invaluable, does have limitations - being resource-intensive and prone to underreporting, particularly for asymptomatic individuals ^15–17^. Such underreporting can significantly skew the accuracy of estimated viral transmissibility. Moreover, these conventional methods may not be effective during the early phase of variant emergence due to the new variant’s low abundance. Wastewater-based surveillance (WBS) has shown promise in monitoring the spread of infectious diseases including the COVID-19 and Mpox ^18–21^. It offers a cost-effective, non-invasive, and near real-time approach to detect the presence of those pathogens within a community and provide early warnings as well as tracking and tracing of outbreaks ^22–29^. However, wastewater is a complex matrix with inhibitors and background nucleic acids ^30,31^, necessitating the development of enhanced tools to boost sensitivity and harness its full potential. Despite the promise of methods like next-generation sequencing, even with amplicon sequencing and target enrichment approaches, detecting emerging variants at trace levels remains a challenge ^32,33^.

To address this gap in sensitivity, we developed an integrated framework including a novel nested allele-specific PCR (NAS-PCR) assay for detecting emerging variants at low prevalence. Using the first identified Omicron variant as a case study, we first employed this NAS-PCR assay with wastewater samples collected from September to December 2021 in Boston to track its early emergence and spread. To validate our wastewater findings that circulation of the Omicron variant predates clinical case reports, we conducted a retrospective analysis of sequenced clinical genomes to trace the global and local emergence and dynamics of Omicron variants. Finally, we quantified the transmissibility by developing a Susceptible-Infected-Viral load model to derive basic reproduction numbers (*R*_0_) from the variant concentrations in wastewater. This integration of molecular diagnostics, wastewater surveillance, and mathematical modeling demonstrates the utility of our framework for early detection and transmissibility assessment of emerging variants.

## 2. Materials and methods

### 2.1. Sample collection and pasteurization

Influent 24-h composite wastewater samples were collected twice a week from September 9 to December 30, 2021, at the Deer Island Wastewater Treatment Plant (DITP) in Massachusetts. There are two influents in the plant, named ABOO and ADOO, serving about 1 million and 1.3 million people in the South and North Boston areas, respectively. Samples (33 from each plant, total 66) were transported to the laboratory on ice for analysis. The average flow rates during the sampling day were obtained from the wastewater treatment plant. In total, 65 composite samples and 1 grab sample (ADOO on 10/5/21) were collected. Samples were first pasteurized at 60 °C for 60 min upon arrival. Next, 50 ml pasteurized samples were passed through a 0.2-µm Steriflip-GP filter (Millipore Sigma) to remove inorganic particles, bacteria, and cell debris. Filtrates were collected to concentrate viral particles. Samples were processed including viral precipitation and extraction during the week they were received.

### 2.2. Viral precipitation, RNA extraction, and RT-qPCR

The filtrate was used for viral enrichment, RNA extraction, and quantitative PCR using methods previously described ^23,36^. In brief, 15 ml filtrate was aliquoted into a 30-kDa Amicon Ultra Centrifugal Filter (Sigma, Cat# UFC9030) and centrifuged at 4,200 rpm for 20 min. The concentrates (about 200 μl) were transferred into a 1.5-ml microtube, and 600 μl Qiagen AVL buffer (Cat# 19073) was added for lysis at room temperature for 10 min. RNA extraction was performed using Qiagen RNeasy kit (Cat# 74182) and eluted with 60-μl nuclease-free H2O. SARS-CoV-2 was tested and quantified using US CDC N1 assay by real-time PCR (Biorad, CFX96 C1000 Touch) using the following program: 50 °C 10 min for reverse transcription, 95 °C 20 s for RT inactivation and initial denaturation, and 48 cycles of denature (95 °C, 1 s) and anneal/extend (60 °C, 30 s). At least two negative controls were included in every PCR run. Pepper mild mottle virus was also measured as an internal reference for sample processing, as previously reported ^23,37^. The remaining volume of each RNA extract was stored at −80 °C. Allele-specific primers and probe were designed to detect the Omicron variant (Om493_498) as previously reported ^38^. Note that minor modifications have been applied to the probe by using a ZEN internal quencher, which showed an improved signal-to-noise ratio and increased sensitivity ^39^.

Cycle threshold (Ct) was converted to viral concentrations (copies per ml) using the standard curves for N1 and Omicron ^37,38^ and further multiplied by the dilution factor (total volume of RNA sample/starting volume of filtered wastewater sample). Two technical replicates were performed for each primer set, and the average of viral concentrations was reported. For ease of visualization, PCR non-detects were assigned the value of 1 in Fig. 1.

**Figure 1.**
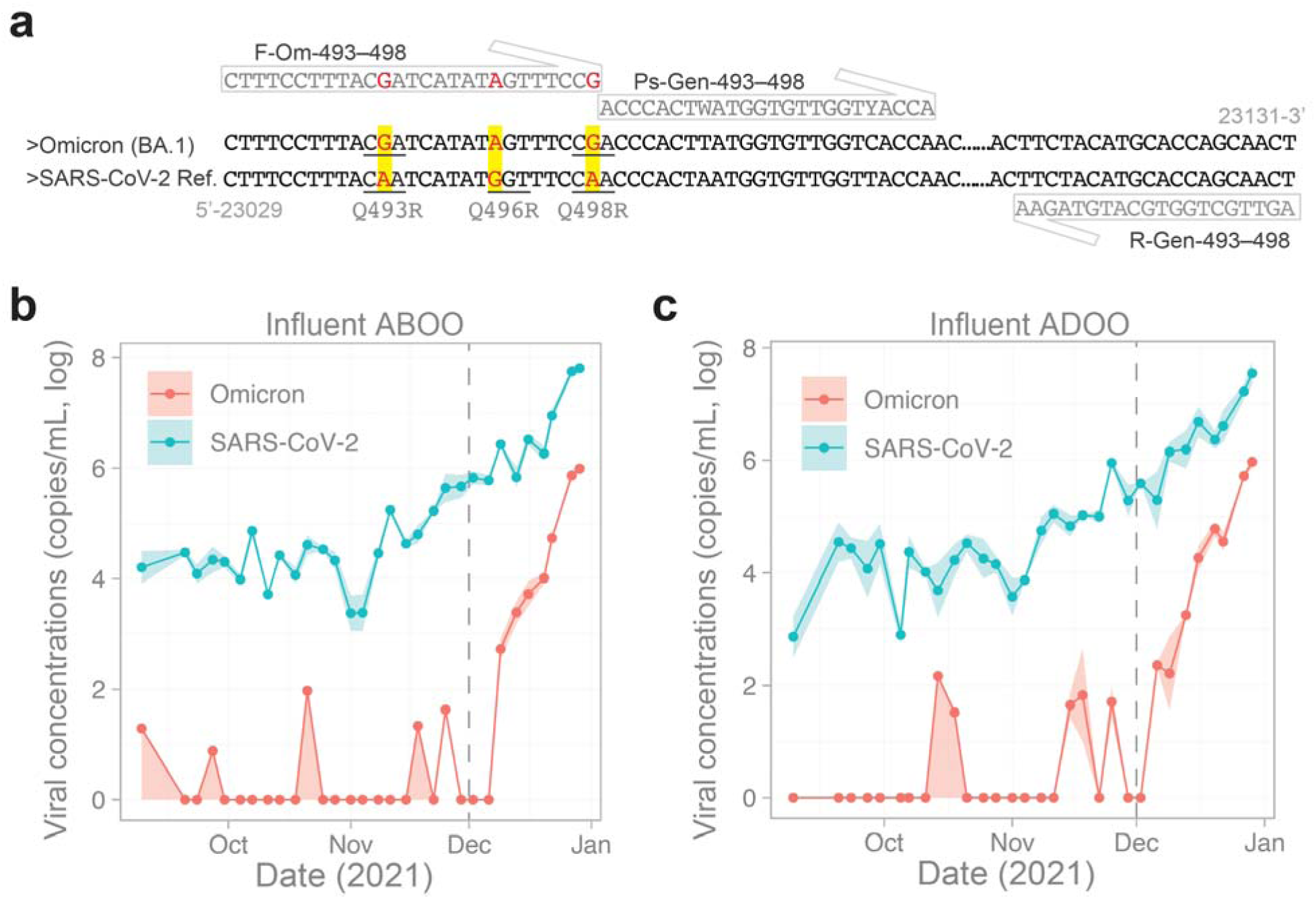
Tracking and quantification of SARS-CoV-2 and Omicron variant in wastewater from September 9 to December 29, 2021. **(a)** Sequence of Omicron variant and SARS-CoV-2 covering the Q493R and Q498R mutations. Allele specific primer and probe sequence and binding sites were shown. ‘SARS-CoV-2 Ref.’ represents the reference genome NC_045512.2. (**b-c**) Omicron and SARS-CoV-2 concentrations in wastewater from September to December 2021 for influents ABOO and ADOO. Solid lines are the mean value and shading area is the maximum and minimum value of the replicates. Negative detection is 1 for visualization purpose. The grey dashed vertical line is 12/01/2021, when the first Omicron infection was announced in the U.S.

### 2.3. Nested allele-specific real-time PCR (NAS-PCR)

NAS-PCR was performed in two steps. First, a 456∼591 bp fragment covering mutations Q493R and Q498R in the spike gene was amplified. Four primers were designed including Forward_aa409 (F409), Forward_aa430 (F430), Reverse_aa581 (R581), and Reverse_aa626 (R626) in **Table S1**. The reference genome for primer design is NC_045512.2. The specificity for primer pairs (F409 and R581; F430 and R581, F430 and R626) was investigated using Primer-BLAST (https://www.ncbi.nlm.nih.gov), in particular against the Refseq mRNA database and Refseq RNA database for *Homo sapiens*, bacteria, and plant organisms. No target templates for the three primer pairs were found in those databases, except for SARS-CoV-2. The three primer pairs were used to detect SARS-CoV-2 with Luna universal real-time PCR master mix (New England Biolabs) using cDNAs of four SARS-CoV-2 positive wastewater samples. RNA was reverse transcribed to cDNA using random primers and ProtoScript IV Reverse Transcriptase (Thermofisher Scientific). Primer pair F409_R581 gave the lowest Ct values across the four wastewater samples and thus was chosen for the NAS-PCR (**Fig. S1**).

The first round of PCR amplification was performed using the Luna universal real-time PCR master mix for all wastewater samples (except ABOO on September 30 and October 5 due to insufficient RNA) using F409_R581 primer pair (519 bp) with the following program: 55 °C and 60 min for reverse transcription; initial denaturation (95 °C, 3 min); 35 cycles of denaturation (95 °C, 10 s), annealing (55 °C, 10 s), and extension (72 °C, 30 s); and a final extension (72 °C, 5 min). Then 4 μl of PCR products were used as DNA template for real-time PCR using the modified Omicron assay (Om493_498) with the above real-time PCR program with F409_R581 pairs without the reverse transcription step. Concurrently, two no-template controls were included for each setup and none of them amplified. Two technical replicates were performed for the real-time PCR reaction.

For NAS-PCR positive samples, we sent PCR products for Sanger sequencing using the primer R-Gen-493-498 (**Table S1**). Sequencing results were then aligned using the NCBI Blastn service (https://blast.ncbi.nlm.nih.gov) with the Standard databases (nr tec.), which outputted 100 sequences producing the most significant alignments and their accession numbers. The aligned percentage of identity ranges from 91.1% to 98.5%. Alignment between the SARS-CoV-2 reference genome (NC_045512.2) and sample sequences was performed using SnapGene software (version 6.1.0). The phylogenetic tree is generated by nextstrain.org ^40^.

### 2.4. Clinical genome data and analysis

Complete SARS-CoV-2 genomes and associated metadata were downloaded from Global Initiative on Sharing All Influenza Data (GISAID). The metadata includes ‘Virus name’, ‘Accession ID’, ‘Collection date’, ‘Submission date’, ‘Location’, and ‘Lineage’. For the epidemiological analysis of Omicron (diversity of subvariants and geographic locations), we only downloaded complete Omicron genomes (with low coverage excluded) collected before November 1, 2021, before Omicron was first reported in South Africa on November 9. As of August 10, 2023, there were 172 Omicron genomes sampled before November 1, 2021. We noticed 11 genomes with incomplete sample collection dates including 8 samples labeled as 01/2021, 1 sample labelled as 2020, 1 sample labelled as 12/2020, and 1 sample labelled as 08/2021. The dates of those sequences were manually changed to 1/31/2021, 12/31/2020, 12/31/2020, and 8/31/2021, respectively, for plotting purposes. The variants were assigned by Pango v.4.3 by GISAID. For the temporal analysis of Omicron emergence in Massachusetts, we collected all the complete genomes (low coverage genomes were excluded, in total 40,152) in the state from 11/15/2021 to 01/06/2022. Genomes with lineages labelled as or starting with “BA.1” (including all of its sub-lineages) and “B.1.1.529” were aggregated as “BA.1”, and the percentages of BA.1 genome were computed by using the total BA.1 genomes divided by the total genomes sampled each day.

### 2.5. Mathematical modeling

Similar to Phan et al. ^41^ and Pell et al. ^42^, we introduce an SI-V (Susceptible-Infected-Viral load) model with one strain of virus (Omicron):

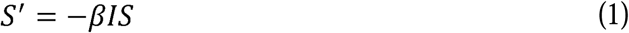

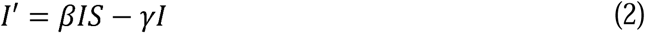

Here, *S* is the initial susceptible population and *I* is the population infected with variant Omicron. The recovered individuals (*R*) do not contribute to transmission dynamics; hence they are omitted. Individuals in the susceptible class are infected upon contact with an infected individual at rate *β*. Infected individuals are assumed to be infectious and stay in the infected class for an average duration of 1/*γ*. The basic reproduction number is given by: 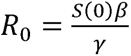. The model is connected to the viral concentration in wastewater by assuming viral shedding is proportional to the infected population ^41,42^.

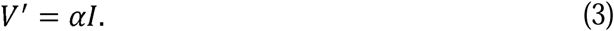

The infected population sheds virus at a composite rate α, which accounts for the average fecal load, average viral shedding rate, and viral loss in the sewer. In Phan et al. (2023), the authors used temporal variation in temperature and segmentation of a population-level viral shedding profile to constrain the individual components of the composite parameter α. However, since we are only interested in a period of several weeks, we directly estimate α to bypass any parameter identifiability issue. Hence, α represents the viral load shed by a single infectious individual per day that remains measurable at the time of sampling. We remark that this model is intended to capture the exponential growth phase following the emergence of a variant (Omicron), so we do not include demography as a parameter. Additionally, the inclusion of an exposed phase (e.g., a brief period when a person is infected but not yet infectious) and a recovered phase is not necessary ^43^ and does not alter the formula for *R*_0_ without demography ^44^. Lastly, the variable *S* in our model does not represent the entire population in the city. When a variant first emerges, the transmission is constrained by the contact network, so it takes time to spread throughout the city. This means that the initial susceptible population (within the first several weeks) refers to the location of the first infections. For this reason, we use various initial susceptible populations (relative to the total population) to provide more robust estimates of *R*_0_. Parameters in the model were provided in **Tables S2∼S4**.

We fit the SI-V model to viral RNA copies in wastewater data by comparing the difference of *V* in every 24-h period, via the minimization of the sum of squared errors (SSE) between the model and data via ^41^:

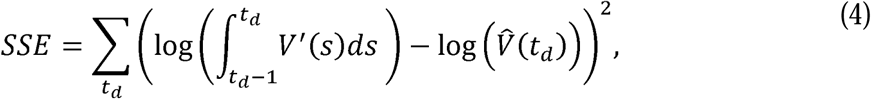

where 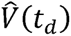 is the total virus in wastewater experimentally measured on day *t_d_*. The minimization is performed using MATLAB built-in functions *fmincon* and *multistart*. For simplicity, we use the approximation 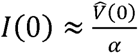 to infer the initial infected population. Following model fitting, we use the best-fit parameters to estimate *R*_0_.

## Results

### Tracking the first emergence of the Omicron variant in wastewater

The first case infected with the SARS-CoV-2 Omicron variant in the United States was announced in California on December 1, 2021. However, due to the limitations of clinical surveillance, it is likely that the circulation of the Omicron variant might have been underway before this official identification. To investigate this possibility, we tested the presence of the Omicron variant by allele-specific real-time PCR in wastewater samples collected bi-weekly from the DITP, spanning from early September 2021 to the end of December 2021. The specificity and sensitivity of this assay, targeting the Omicron signature mutations Q493R to Q498R in the spike gene for both BA.1 and BA.2 lineages (**Fig. 1a**), were validated in our previous study ^38^.

The Omicron variant and SARS-CoV-2 were quantified in wastewater samples. Results showed that the concentration of SARS-CoV-2 was relatively stable from September to early November, followed by a ∼50-fold increase in December in both DITP influents (**Fig. 1b-c**). Consistent detections of Omicron (following exponential growth) started on December 6 in ADOO, and on December 9, 2021 in ABOO influent wastewater. Sporadic detections of the Omicron variant were observed in samples from ABOO influent including two samples in September (Sep. 9 and 27), one sample on October 21, and two samples in November (Nov. 11 and 25). Omicron was also found in influent samples from ADOO, including two samples in October (Oct. 14 and 18), and three samples in November (Nov. 15, 18, and 25).

### The Omicron variant is detected in wastewater in September 2021 by NAS-PCR

Sporadic positive detections were also observed in the months before December (**Fig. 1**). To verify this and enhance the sensitivity of detection, we developed a nested allele-specific real-time PCR (NAS-PCR) for Omicron in wastewater samples (**Fig. 2a**). The NAS-PCR was developed first by designing three primer pairs (F409/R581, F430/R581, and F430/R626) to amplify a spike gene fragment from aa409 to aa626, which covers the Q493R and Q498R mutations in the Omicron variant. Based on four SARS-CoV-2 positive wastewater samples, we found that F409/R581 had the lowest Ct values compared to the other two primer pairs (**Fig. S1**), and thus we chose it for the initial amplification. Next, the F409/R581 PCR product (519 bp) was used as template for the allele-specific real-time PCR ^38^ for Omicron detection.

**Figure 2.**
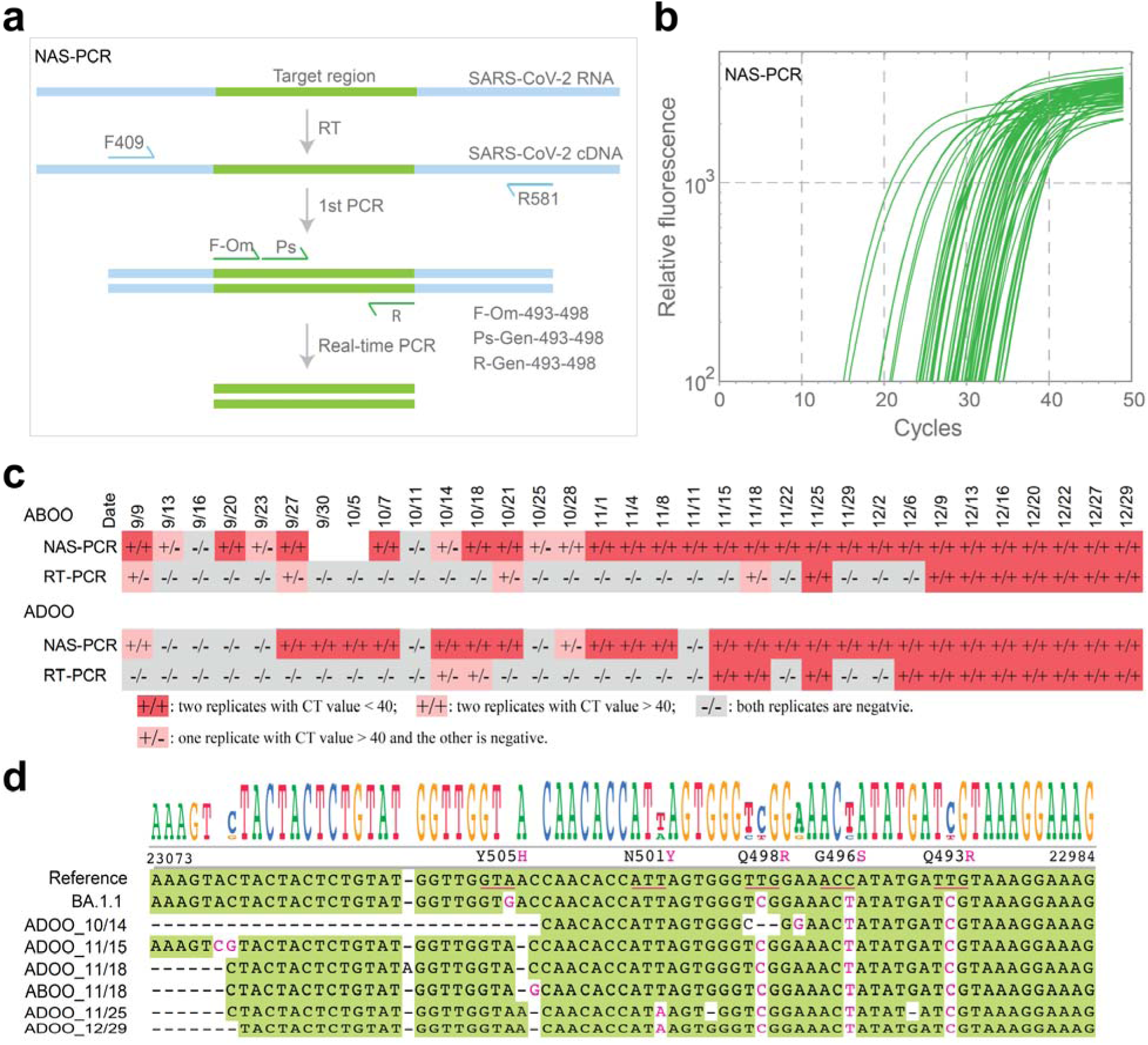
Nested real-time PCR detects presence of the Omicron variant in community wastewater from early September 2021. **(a)** Illustration of NAS-PCR for Omicron detection. F409, F430, R581, and R626 are the primers designed for the first round of amplification to include the Q493R and Q498R region. **(b)** NAS-PCR amplification curves from all tested wastewater samples, dated from 9/9/2021 to 12/29/2021, displaying only samples with Ct value < 40. **(c)** Summary of the detection results using NAS-PCR and RT-PCR for ABOO (top) and ADOO (bottom). Detections with both replicates are positive and Ct < 40 are highlighted in red; plausible detections with Ct value > 40 are highlighted in pink; and negative detections are colored grey. **(d)** Sequence alignment results from Sanger sequencing for 6 of the positive wastewater samples derived by NAS-PCR. The reference genome is SARS-CoV-2 isolate Wuhan-Hu-1 NC_045512.2, and the accession number for BA.1.1 is ON042406. Sequenced samples were named (on the left) using the format: sewershed_sampling date.

NAS-PCR results show that the earliest positive detection of Omicron occurred on September 9, 2021. Specifically, Omicron was first detected in ABOO samples on September 9, 20, and 27, October 7, 18, 21, and consistently from November 1 (**Fig. 2b-c**). For ADOO, the first positive detection was observed on September 27, while indeterminate detection (Ct value > 40) started on September 9. Overall, Omicron was detected by NAS-PCR in 24 out of 31 samples (77.4%) from ABOO and 24 out of 33 samples (72.7%) from ADOO, compared to 8/33 (24.2%) in ABOO and 11/33 (33.3%) in ADOO using the non-nested real-time PCR assay, respectively (**Fig. 2b-c**). Among the 24 positive NAS-PCR detections, 21 samples (87.5%) were concurrently positive in both influents on the same sampling day. To further confirm the NAS-PCR results, we Sanger sequenced the NAS-PCR products and aligned the sequences against the NCBI database. Sequencing results confirmed a 91.1% ∼ 98.5% match in identity to the SARS-CoV-2 BA.1 spike gene (**Fig. 2d**). The results combined with the NAS-PCR detections support the conclusion that the Omicron variant was circulating in the community months before the first clinical case report in the country on December 1, 2021.

### Dating the emergence of the Omicron variant from global clinical sequencing data

Given the delay between clinical sample collection, genome sequencing, and data submission, we investigated the emergence of Omicron from the clinical sequencing dataset in GISAID. We compiled the complete genomes of Omicron BA.1 and its sub-lineages, which were sampled before 11/1/2021, prior to the first Omicron case being reported in the world. In total, 172 Omicron genomes including BA.1, BA.2, BA.4, and BA.5 and their sub-lineages were found, with 5 genomes collected in 2020 and 167 in 2021 (**Fig. 3a**). Of the 172 Omicron genomes, BA.1.1, BA.1, BA.2, and BA.1.15 were the four most abundant lineages in a total of 30 Omicron lineages (**Fig. 3a and Fig. S2a**). The phylogenetic tree also showed the two main clades BA.1 and BA.2, with diverse distributions of those genomes in different continents. (**Fig. S3**). It should be noted that the earliest BA.1 genomes were collected from Asia and Europe, rather than Africa where the first Omicron case was reported (**Fig. 3b**). In total, 89, 48, and 19 genomes were collected from Europe, North America, and Africa, respectively (**Fig. S2b**). These results indicated that global circulation of Omicron infections might have occurred much earlier than when it was first flagged in early November 2021, aligning with and corroborating our findings from the NAS-PCR wastewater analysis.

**Figure 3.**
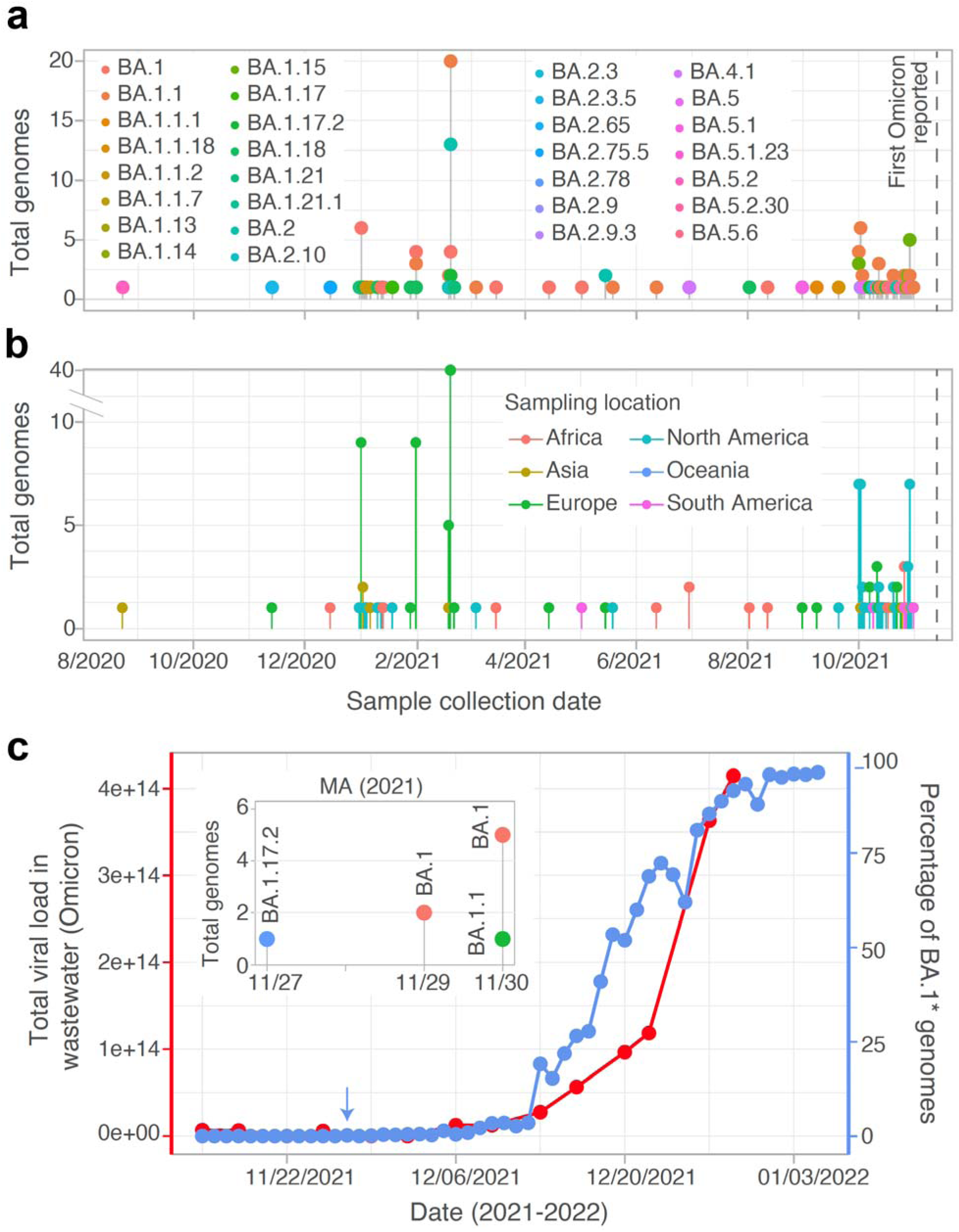
Analysis of clinical Omicron genomes and comparison with wastewater viral loads. **(a-b)** The number of complete SARS-CoV-2 Omicron variant genomes collected before 11/1/2021, colored by lineages (**a**) and sampling locations (**b**). The grey dotted lines represent the date of the first Omicron case reported in the world (11/9/2021). (**c**) Comparison of Omicron viral load in wastewater (red) and percentage of Omicron BA.1 genomes in MA (blue). Insert: Omicron lineages in MA before 12/1/2021. BA.1*: include BA.1 and its sub-lineages as well as B.1.1.529. The blue arrow represents the first date for which Omicron was tested positive via clinical sequencing data. Total viral load equals to viral concentrations in wastewater multiplied by the daily flow volume.

We next analyzed the emergence of Omicron in Massachusetts. The first clinical Omicron genome in MA is BA.1.17.2, sampled on 11/27/2021 (**Fig. 3c**, insert), in contrast to the first Omicron-positive wastewater sample collected on 9/9/2021 by NAS-PCR. Wastewater testing could have conferred a lead time of more than two months compared to clinical testing. We then plotted the growth of Omicron cases using the percentage of clinically reported BA.1 genomes in MA against the wastewater viral load data (Omicron concentration multiplied by flow volume). As shown in **Fig. 3c**, both datasets include an exponential growth period starting from mid-December, but the trend of BA.1 growth based on clinical genomes precedes the measured wastewater data.

### Estimated transmissibility of SARS-CoV-2 Omicron variant from wastewater data by an SI-V model

In our previous work, we developed a Susceptible-Exposed-Infectious-Recovered-and-Viral load model to estimate COVID-19 prevalence and predict SARS-CoV-2 transmission dynamics using the temporal wastewater data ^41,42^. We set out to fit a simplified Susceptible-Infected-Viral load (SI-V) model with measured Omicron data to estimate the basic reproduction number (*R*_0_), which is a fundamental gauge of pathogens’ transmissibility in the population ^34,35,45,46^. As shown in **Fig. 1**, Omicron has been continually detected and quantified beginning on December 9 in influent at ABOO and December 6 at ADOO, so to ensure the quality of model fitting, we used the wastewater data from December 9 to fit the SI-V model for both ABOO and ADOO data. Results in **Fig. 4a-b** showed that the model recapitulated the growing dynamics of Omicron’s viral load in both the ABOO and ADOO sewersheds. With this model, we identified two key parameters, the infection rate *β* and the average duration of infectiousness 1/γ, in the sewershed for *R*_0_ estimation.

**Figure 4.**
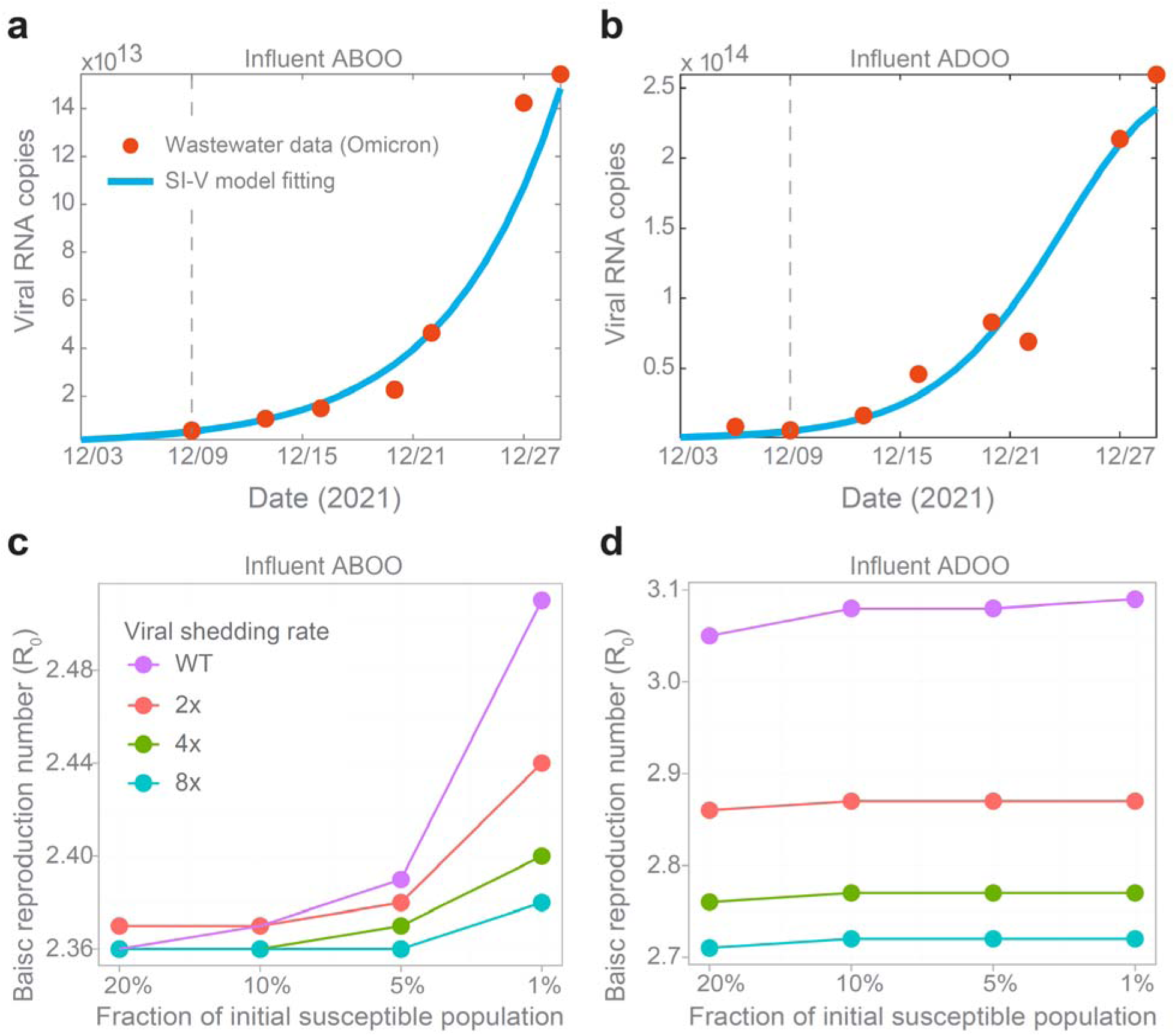
Estimated transmissibility of the SARS-CoV-2 Omicron variant from wastewater data. **(a-b)** SI-V model fitting using 7 data points measured from ABOO and ADOO influents starting from December 9, 2021 (as indicated by the dashed grey line). The y-axis is the total viral load, i.e., viral concentrations multiplied by flow volume on the sampling day. **(c-d)** Estimates of *R*_0_ with varying percentages of initial susceptible population and viral shedding rates. WT: fitted baseline viral shedding rate for wild-type SARS-CoV-2 ^53^; and 2x∼8x: viral shedding rate with two to eight fold of the wild-type strain.

*R*_0_ is influenced by the size of the initial susceptible population *S*(0), the exact value of which is challenging to quantify. We thus tested the effect of changing the size of the initial susceptible population between 20% and 1% (as fraction of the total population in the Boston area) on the estimate of *R*_0_. Results showed that *R*_0_ slightly increased from 2.36 to 2.51 for an increasing *S*(0) for sewershed ABOO (**Fig. 4c**, purple line), and varied between 3.05 and 3.09 for ADOO (**Fig. 4d**, purple line). These *R*_0_ estimates are consistent with other estimates reported in the literature, ranging from 1.5 to roughly 10 ^47–49^. In particular, an earlier study that estimated the effective reproduction number of Omicron gives estimates to be around 2 to 3 in December 2021 and also showed significant temporal variation in the estimates of the reproductive number for Omicron over time ^48^. It is well known that estimation based on limited data during the early phase of an outbreak may result in a large uncertainty of estimates of *R*_0_ ^50–52^. Thus, to further test whether estimates of *R*_0_ are impacted by the number of used data points ^51^, we fit the model with 6 data points starting from December 13 (**Fig. S4**) and found a small deviation compared to the *R*_0_ estimated using 7 data points. Specifically, *R*_0_ ranged from 2.41∼2.61 for ABOO (2.1%∼3.9% difference to the estimate using 7 data points), and 2.84∼2.90 (6.0%∼7.1% difference to the estimate using 7 data points) for ADOO (**Fig. S5**). These results showed that *R*_0_ inferred from wastewater data is relatively stable in terms of initial susceptible population size and model fitting.

The viral shedding rate may impact SI-V modelling results. To further investigate the extent to which viral shedding rate affects the estimation of *R*_0_, we tested the model with different viral shedding rates, i.e., 2x, 4x, and 8x fold of the fitted baseline composite shedding rate α ^53^, since the exact shedding for Omicron at the population level is not known. The results showed a slight reduction in the basic reproduction number, up to 11.9% (2.51 to 2.38 for ABOO; 3.09 to 2.71 for ADOO, when the viral shedding rate was increased by a factor of 2 to 8 (**Figure 4c-d**). Similar results were also observed using 6 data points for model fitting (**Fig. S5b**). Altogether, these analyses suggest that the SI-V model, applied to wastewater surveillance data, could be used to infer the transmission reproduction number *R*_0_. The *R*_0_, estimated to be 2 to 3 for Omicron BA.1 during the first Omicron wave in the Boston area, is relatively stable across different scenarios including initial susceptible population size, the number of data points for model fitting, and viral shedding rates.

## Discussion

In this study, we aimed to develop a framework for wastewater-based variant surveillance by tracking the emergence of the first SARS-CoV-2 Omicron variant in a population using a novel method NAS-PCR and quantifying the variant’s transmissibility using a compartmental epidemic model SI-V on the wastewater viral load of Omicron. The NAS-PCR offers a sensitive and cost-efficient approach for early detection of a specific SARS-CoV-2 variant at low prevalence, which can easily be implemented into existing molecular workflows to quantify the virus in clinical or environmental samples. The SI-V model bridges wastewater surveillance data with epidemiological modelling, providing a quantitative understanding of the Omicron’s capability for transmission within a sewershed. The framework is thus useful for understanding the emergence and pandemic potential of pathogens during the early phase of an outbreak.

NAS-PCR integrates nested PCR and allele-specific PCR for improved sensitivity and mutation-specific detection. Nested PCR has long been used for detecting pathogens with low concentrations owing to its higher sensitivity and specificity, compared to the conventional PCR ^54–58^. Sirakov and colleagues’ nested PCR showed 100% sensitivity and specificity for SARS-CoV-2 detection in cats ^59^. La Rosa et al. (2021) developed nested RT-PCR assays for the detection of mutations in the spike gene of SARS-CoV-2 ^55^. However, nested, allele-specific PCR is a novel approach that has not been used for detecting viral variant in wastewater. While one potential concern of nested PCR is cross-contamination during the transfer of the first-step PCR product for the second amplification step, the presence of negative amplifications in some samples (2 out of 31 in ABOO and 7 out of 33 for ADOO in **Fig. 2c**), as well as the non-amplification in no-template controls, suggest this risk is low. The presence of Omicron in wastewater samples collected in October and November was further confirmed by Sanger sequencing. The nucleotide variations in the sequencing results of NAS-PCR products (**Fig. 2d**) further support the minimal likelihood of contamination. The NAS-PCR method serves as a valuable complement to genomic surveillance, facilitating early detection and tracking of new SARS-CoV-2 variants when their prevalence is low.

A retrospective analysis of genomes deposited in the GISAID database uncovered 172 complete Omicron genomes sequenced from clinical samples collected before 11/1/2021. Among these, 5 genomes were from 2020, and 167 were from 2021 (**Fig. 3a**). The first identified Omicron genome was in Japan on 8/23/2020, over a year before the first officially reported Omicron case on 11/9/2021. These genomes were collected from 30 countries in 6 continents, corroborating the NAS-PCR findings (**Fig. 2**) and implying the global presence of the Omicron variant months before the first massive Omicron variant wave. However, all 172 genomes were submitted after December 11, 2021, following the first officially reported case in November 2021. This discrepancy raises two potential issues: 1) possible errors in the metadata ^60^, particularly the sample collection dates, although 161 of the 172 genomes have complete date information; and 2) potential sample contamination by post-November 2021 Omicron BA.1 samples during processing or sequencing, noting that precise extraction and sequencing dates are unavailable. Notably, these genomes were originally sampled from 71 different laboratories, which could lend credibility to the findings by reducing the likelihood of a single source error. However, it also introduces complexity in validating the data across diverse laboratory protocols and standards. While we present these genomic analysis results to provide a comprehensive view of available data and its alignment with the wastewater findings, we acknowledge the limitations in definitively addressing the two concerns within the scope of this study. Therefore, we present this information transparently, allowing readers to critically evaluate the implications of these findings and their potential to reshape our understanding of variant’s emergence (Omicron) and early spread.

Historically, the epidemic peak of SARS-CoV in southeast China was in early February 2003. However, sporadic cases (ranging from 1 to 9) were retrospectively identified in samples from 11/16/2002 in multiple satellite cities of Guangdong Province ^11,12^. This implies that SARS-CoV was already circulating within the population for over two months before triggering a significant regional outbreak. This historical example suggests the possibility that coronaviruses can circulate within a population for months, or even longer, prior to sparking a pandemic. The observation not only prompts curiosity about the underlying reasons for its circulation but also emphasizes the importance of tools for tracking and understanding the transmissibility of viruses whilst at trace levels as a proactive measure to prevent potential large-scale outbreaks.

Wastewater-based surveillance (WBS) has been widely implemented in many countries and recommended by (non-) governmental bodies including the World Bank, the US-CDC, and the European Commission. However, the translation of WBS data for epidemiological inference remains largely unexplored. The SI-V model helps bridge wastewater surveillance data and epidemiological models, providing a quantitative understanding of Omicron’s capability for transmission within a sewershed. In our previous work, we developed an SEIR-V model to integrate WBS data into a compartmental epidemic model and demonstrated that the model effectively recapitulated the temporal dynamics of viral load in Boston wastewater ^41^. Based on the present model, we showed that Omicron concentrations in wastewater can be further used to estimate the basic reproduction number *R*_0_, which describes the average number of secondary cases caused by a single infectious person in a completely susceptible population. *R*_0_ is typically estimated based on contact tracing data or clinical data such as reported incidence, deaths, and hospitalizations ^44,61,62^, however, those data may be limited by underreporting of asymptomatic cases in the population, leading to an inaccurate *R*_0_ estimate. As such, using the population-level wastewater surveillance data may provide a relatively less biased estimate of the viral transmissibility. Our model inferred the *R*_0_ is 2∼3 during the early Omicron outbreak in the Great Boston area, which agrees with the reported range of *R*_0_ ^47–49^. We further showed that this estimate is robust vis-à-vis changes in the initial population size, data points for model fitting, and viral shedding rates (see Fig. 4). While empirical estimation of the reproductive number *R*_e_ from wastewater data has been previously reported using statistical tools like EpiEstim ^63^, to our best knowledge, this is the first study using a mechanistic, dynamic SIR-type model to directly estimate *R*_0_ for Omicron from wastewater surveillance data.

In conclusion, our study presents an integrated framework that combines the nested allele-specific PCR, wastewater-based surveillance, and mechanistic epidemic modeling for variant monitoring. Using the first Omicron BA.1/2 as a case study, we demonstrated that this framework’s capability for early detection and community-level transmissibility assessment. Our findings suggest that the Omicron variant might have been in circulation months before its first clinical detection, highlighting the value of these new methods in early variant detection. This framework’s modular components can be readily adapted to other viruses and their variants, and broaden the wastewater-based surveillance toolkit and strength public health preparedness for future epidemics.

## Data Availability

All data produced in the present work are contained in the manuscript

## Author Contributions

F.W. conceptualized the project, performed the experiments, and clinical genome data analysis. T.P. performed the modeling analysis, with inputs from S.B., B.P., and Y.K. S.F.R and S.P. helped with sample collection and transfer. X.C. and all other authors contributed to the discussion, writing, and editing of the manuscript. E.J.A. and F.W. supervised the project. All authors read and approved the manuscript.

## Code Availability

All data produced in the present work are contained in the manuscript. The code for the mathematical modeling and genomic data analysis will be shared with the paper formal publication.

## Acknowledgements

We gratefully acknowledge all data contributors, i.e., the Authors and their Originating laboratories responsible for obtaining the specimens, and their Submitting laboratories for generating the genetic sequence and metadata and sharing via the GISAID Initiative, on which this research (Fig 3, Fig. S2 and S3) is based. We are grateful to Amy Xiao and Annie Shen for helping to ship the samples. This work is supported by the National Science Foundation (DMS- 2421257) and UT system Rising STARs award. T.P. is supported by Director’s postdoctoral fellowship at Los Alamos National Laboratory. This work was also supported by the MIT Center for Microbiome Informatics and Therapeutics to EJA, the National Research Foundation, Prime Minister’s Office, Singapore, under its Campus for Research Excellence and Technological Enterprise (CREATE) program funding to the Singapore-MIT Alliance for Research and Technology (SMART) Antimicrobial Resistance Interdisciplinary Research Group (AMR IRG), the Intra-CREATE Thematic Grant (Cities) grant NRF2019-THE001-0003a to JT and EJA and funding from the Singapore Ministry of Education and National Research Foundation through an RCE award to Singapore Centre for Environmental Life Sciences Engineering (SCELSE).

---

**Figure S1.**
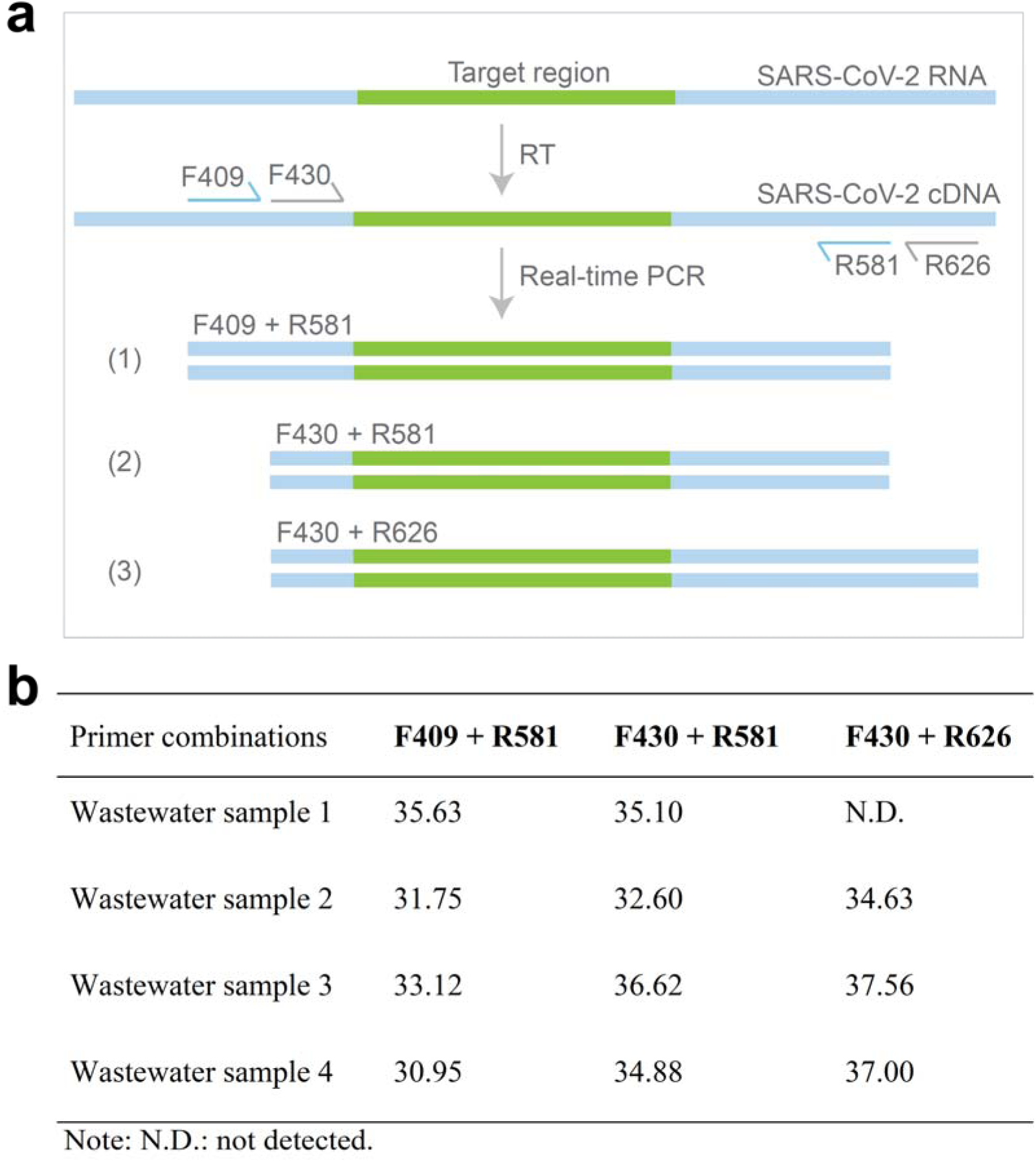
Primer selection for the first step of NAS-PCR. (a) Illustration of PCR strategy to select the primers for the first step of NAS-PCR. Four specific primers F409, F430, R581, and R626 were designed for the first round of amplification to include the Q493R and Q498R region. (b) Performance comparison of primer combinations (Ct value) for the first step of NAS-PCR, tested by four wastewater samples’ RNA. F409+R581 consistently showed the best performance (lowest Ct value) among the three combinations.

**Figure S2.**
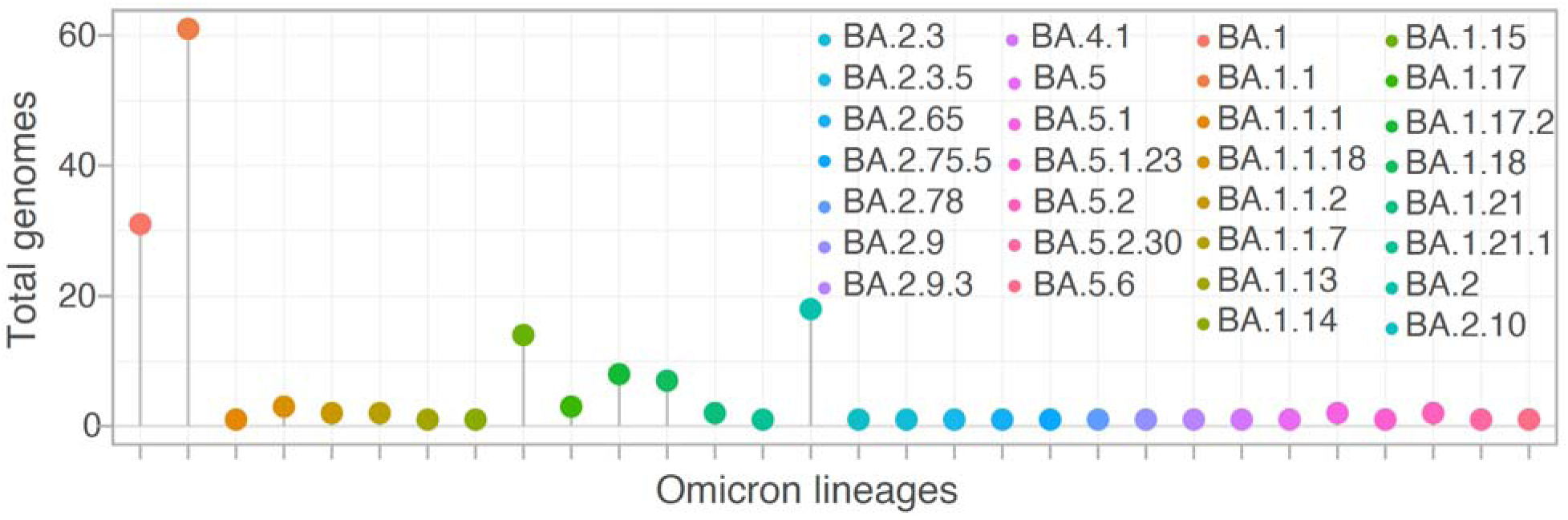
The number of Omicron lineage genomes for samples collected before the first Omicron case was reported. The number of complete SARS-CoV-2 Omicron variants’ genomes identified in GISAID before 11/1/2021.

**Figure S3.**
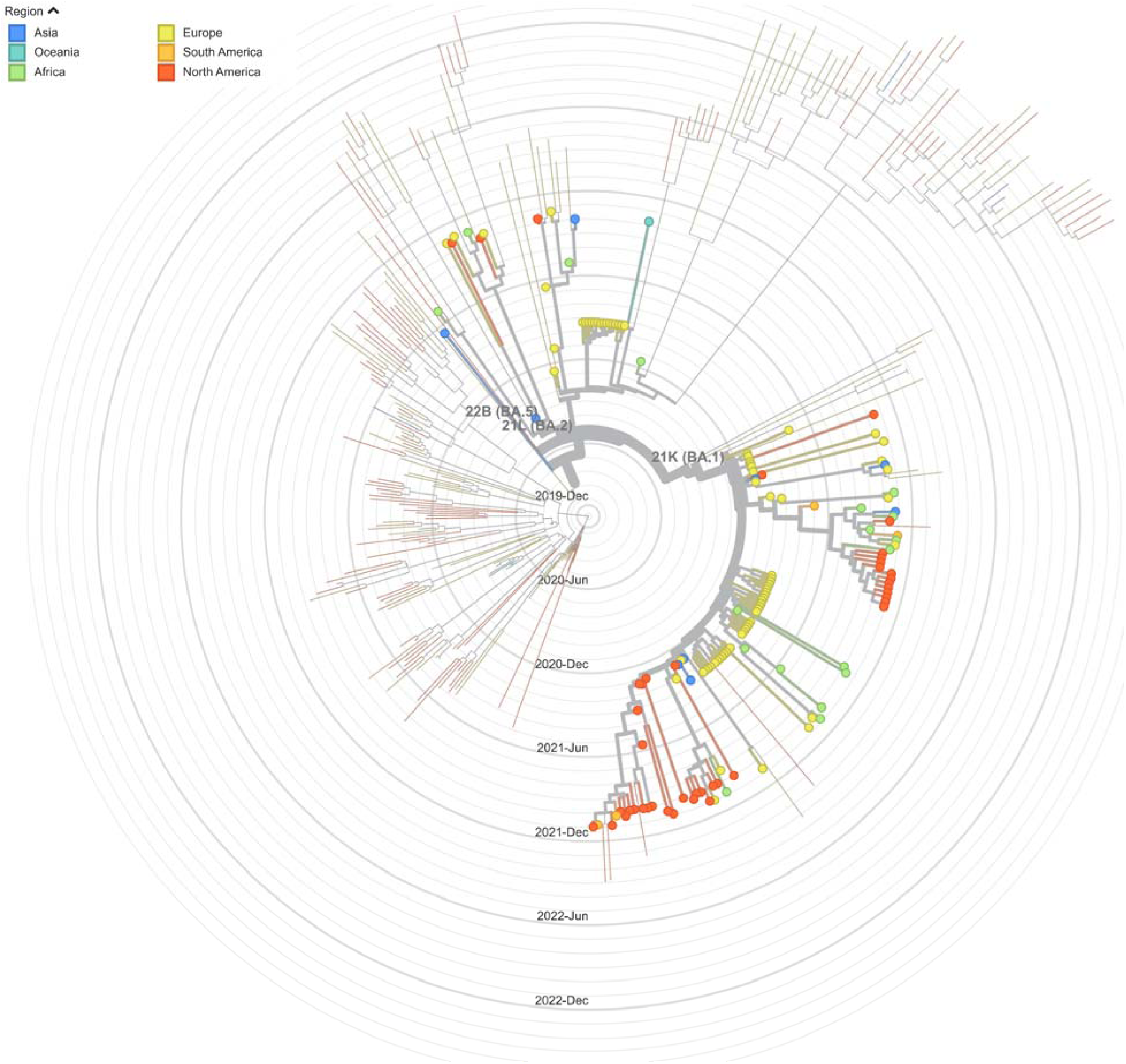
A radial phylogenetic tree of SARS-CoV-2 Omicron variants sampled before November 1, 2021. SARS-CoV-2 complete genomes were obtained from GISAID, and clustered by clade and colored by sampling regions. 151 of 172 samples were displayed in the tree, and the rest 21 samples were dropped during filtering due to insufficient metadata information. Background layout (grey rings) represents sampling date and thin lines are branches for reference genomes of SARS-CoV-2 and variants generated by nextstrain.org.

**Figure S4:**
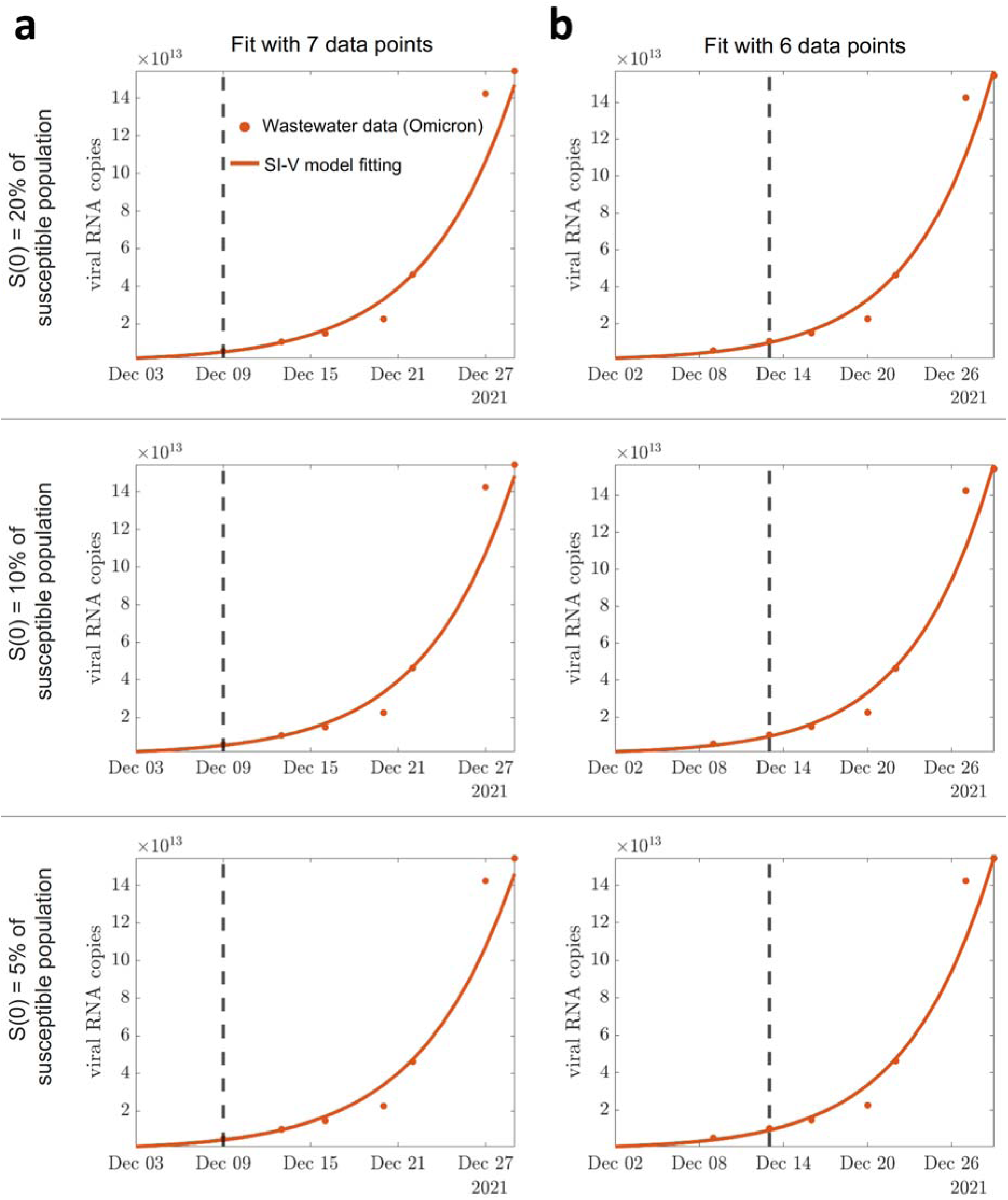
SI-V model fitting using wastewater data measured in influent ABOO. The best fit to the total viral load of the Omicron variant in wastewater using (**a**) 7 data points versus (**b**) 6 data points. Data on the right of the dashed vertical line are used for the fitting. Top to bottom are fittings with 20%, 10%, and 5% of the total population as the initial susceptible population.

**Figure S5:**
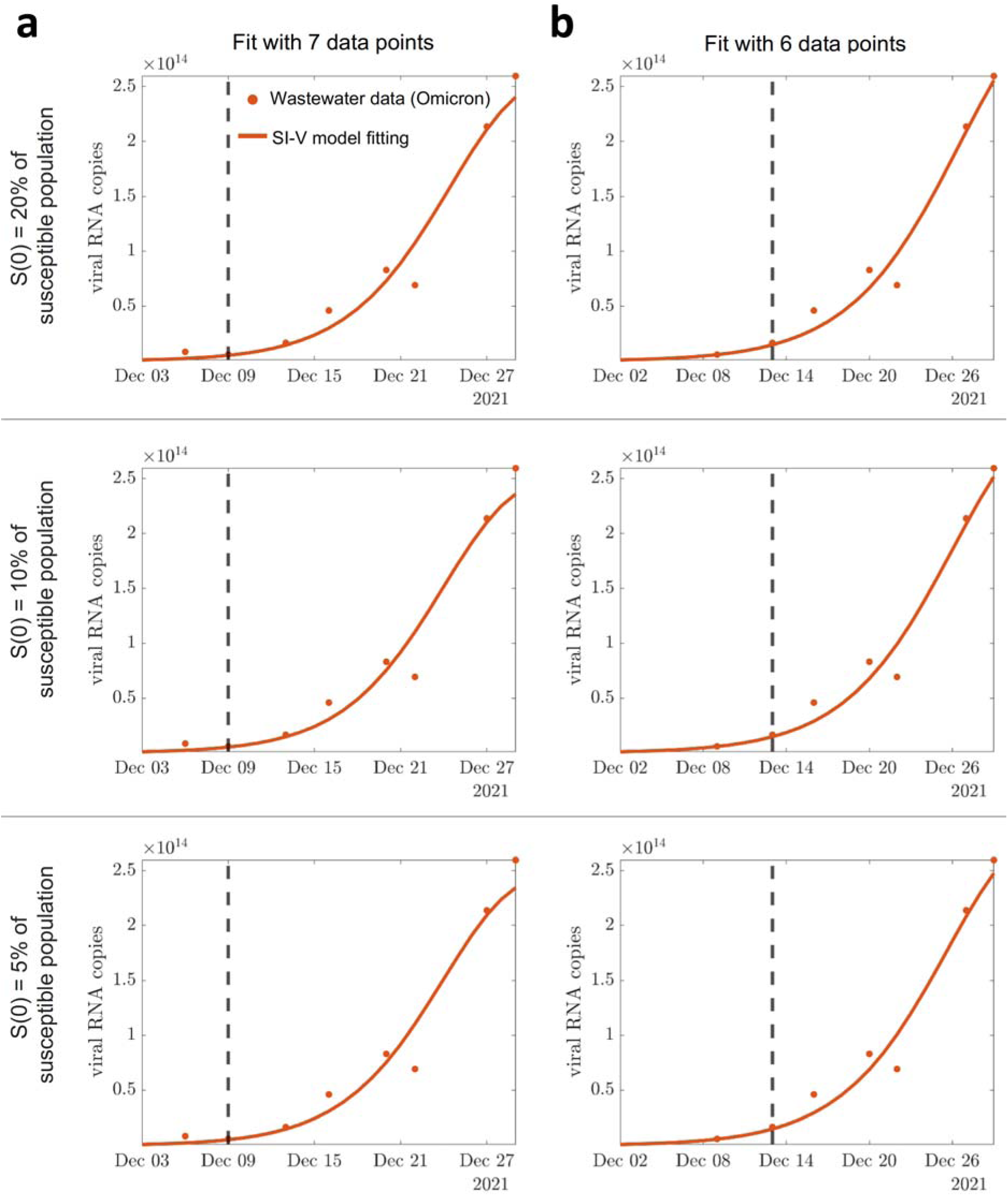
SI-V model fitting using wastewater data measured in influent ADOO. The best fit to the total viral load of the Omicron variant in wastewater using (**a**) 7 data points versus (**b**) 6 data points. Data on the right of the dashed vertical line are used for the fitting. Top to bottom are fittings with 20%, 10%, and 5% of the total population as the initial susceptible population.

**Table S1.**
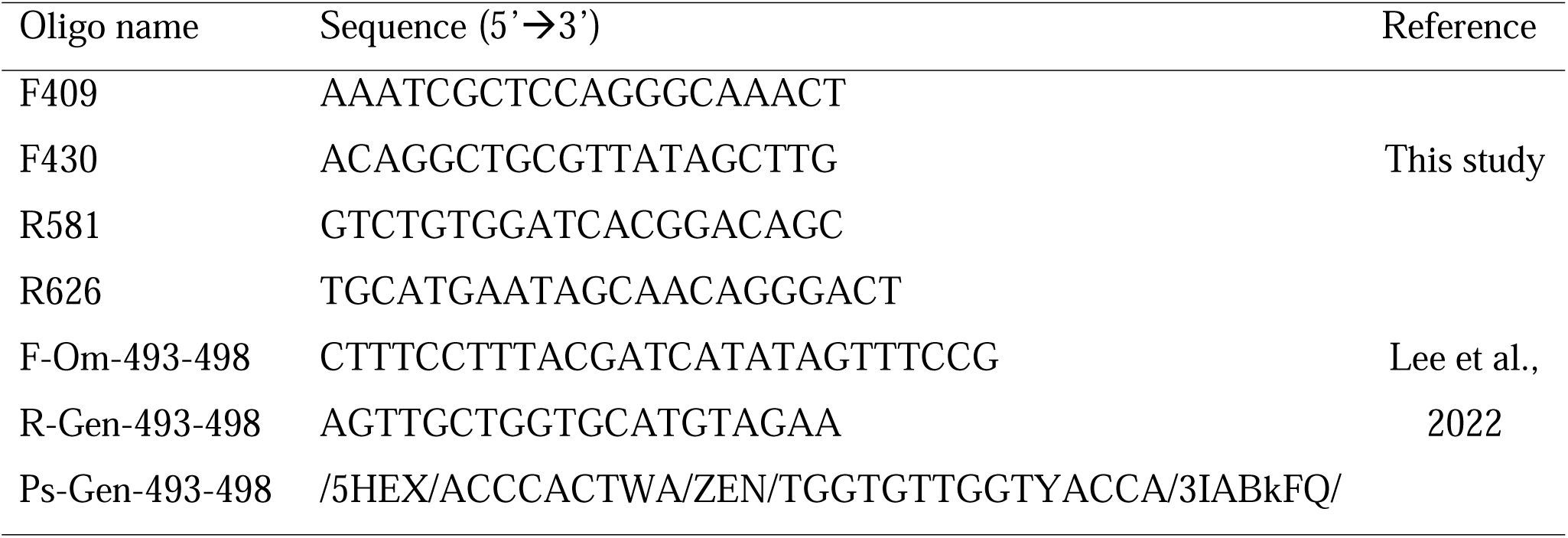
Oligos and sequence in this study.

**Table S2.** Parameters in the SI-V model for ABOO influents.

| ABOO | Baseline | 2X $\alpha$ | 4X $\alpha$ | 6X $\alpha$ | 8X $\alpha$ |
| --- | --- | --- | --- | --- | --- |
| S(0) = 20%, 6 pt | $\beta = 1.51\text{e-}6, \alpha = 6.13\text{e}10$ | $\beta = 1.51\text{e-}6$ | $\beta = 1.50\text{e-}6$ | $\beta = 1.50\text{e-}6$ | $\beta = 1.50\text{e-}6$ |
| S(0) = 10%, 6 pt | $\beta = 3.03\text{e-}6, \alpha = 6.84\text{e}10$ | $\beta = 3.02\text{e-}6$ | $\beta = 3.01\text{e-}6$ | $\beta = 3.01\text{e-}6$ | $\beta = 3.01\text{e-}6$ |
| S(0) = 5%, 6 pt | $\beta = 6.10\text{e-}6, \alpha = 7.79\text{e}10$ | $\beta = 6.06\text{e-}6$ | $\beta = 6.03\text{e-}6$ | $\beta = 6.03\text{e-}6$ | $\beta = 6.02\text{e-}6$ |
| S(0) = 1%, 6 pt | $\beta = 3.27\text{e-}5, \alpha = 7.80\text{e}10$ | $\beta = 3.13\text{e-}5$ | $\beta = 3.07\text{e-}5$ | $\beta = 3.05\text{e-}5$ | $\beta = 3.04\text{e-}5$ |
| S(0) = 20%, 7 pt | $\beta = 1.48\text{e-}6, \alpha = 4.05\text{e}10$ | $\beta = 1.48\text{e-}6$ | $\beta = 1.48\text{e-}6$ | $\beta = 1.48\text{e-}6$ | $\beta = 1.48\text{e-}6$ |
| S(0) = 10%, 7 pt | $\beta = 2.96\text{e-}6, \alpha = 7.48\text{e}10$ | $\beta = 2.96\text{e-}6$ | $\beta = 2.95\text{e-}6$ | $\beta = 2.95\text{e-}6$ | $\beta = 2.95\text{e-}6$ |
| S(0) = 5%, 7 pt | $\beta = 5.98\text{e-}6, \alpha = 6.97\text{e}10$ | $\beta = 5.94\text{e-}6$ | $\beta = 5.92\text{e-}6$ | $\beta = 5.91\text{e-}6$ | $\beta = 5.91\text{e-}6$ |
| S(0) = 1%, 7 pt | $\beta = 3.14\text{e-}5, \alpha = 7.83\text{e}10$ | $\beta = 3.05\text{e-}5$ | $\beta = 3.00\text{e-}5$ | $\beta = 2.98\text{e-}5$ | $\beta = 2.97\text{e-}5$ |

Note: $\alpha$ represents the viral shedding load per day; $\beta$ is the infection rate; and $S$ is the initial susceptible population.

**Table S3.** Parameters in the SI-V model for ADOO (middle) influents.

| ADOO | Baseline | 2X $\alpha$ | 4X $\alpha$ | 6X $\alpha$ | 8X $\alpha$ |
| --- | --- | --- | --- | --- | --- |
| S(0) = 20%, 6 pt | $\beta = 1.36\text{e-}6, \alpha = 4.28\text{e}9$ | $\beta = 1.28\text{e-}6$ | $\beta = 1.24\text{e-}6$ | $\beta = 1.23\text{e-}6$ | $\beta = 1.22\text{e-}6$ |
| S(0) = 10%, 6 pt | $\beta = 2.76\text{e-}6, \alpha = 7.95\text{e}9$ | $\beta = 2.57\text{e-}6$ | $\beta = 2.49\text{e-}6$ | $\beta = 2.46\text{e-}6$ | $\beta = 2.45\text{e-}6$ |
| S(0) = 5%, 6 pt | $\beta = 5.56\text{e-}6, \alpha = 1.52\text{e}10$ | $\beta = 5.17\text{e-}6$ | $\beta = 4.99\text{e-}6$ | $\beta = 4.93\text{e-}6$ | $\beta = 4.90\text{e-}6$ |
| S(0) = 1%, 6 pt | $\beta = 2.79\text{e-}5, \alpha = 7.50\text{e}10$ | $\beta = 2.58\text{e-}5$ | $\beta = 2.49\text{e-}5$ | $\beta = 2.47\text{e-}5$ | $\beta = 2.45\text{e-}5$ |
| S(0) = 20%, 7 pt | $\beta = 1.45\text{e-}6, \alpha = 3.54\text{e}9$ | $\beta = 1.37\text{e-}6$ | $\beta = 1.33\text{e-}6$ | $\beta = 1.31\text{e-}6$ | $\beta = 1.30\text{e-}6$ |
| S(0) = 10%, 7 pt | $\beta = 2.97\text{e-}6, \alpha = 6.03\text{e}9$ | $\beta = 2.76\text{e-}6$ | $\beta = 2.66\text{e-}6$ | $\beta = 2.63\text{e-}6$ | $\beta = 2.61\text{e-}6$ |
| S(0) = 5%, 7 pt | $\beta = 5.94\text{e-}6, \alpha = 1.20\text{e}10$ | $\beta = 5.53\text{e-}6$ | $\beta = 5.32\text{e-}6$ | $\beta = 5.26\text{e-}6$ | $\beta = 5.23\text{e-}6$ |
| S(0) = 1%, 7 pt | $\beta = 2.97\text{e-}5, \alpha = 5.98\text{e}10$ | $\beta = 2.76\text{e-}5$ | $\beta = 2.66\text{e-}5$ | $\beta = 2.63\text{e-}5$ | $\beta = 2.61\text{e-}5$ |

Note: $\alpha$ represents the viral shedding load per day; $\beta$ is the infection rate; and $S$ is the initial susceptible population.

**Table S4.** Other parameters in the SI-V model.

| Other parameters | Value | References |
| --- | --- | --- |
| Susceptible population ($S$) | 1000000 (ABOO) | Wu et al.,<br>2022 |
|  | 1300000 (ADOO) |  |
| Infection duration ($1/\gamma$) | 8 days | Killingley et al. 2022, Phan et al. 2023 |

